# Genetic consequences of serial sperm donation

**DOI:** 10.1101/2025.09.19.25336188

**Authors:** Thomas M. Zheng, Alejandro Mejia-García, Claude Bhérer, Catherine Laprise, Anne-Marie Laberge, Simon Gravel

## Abstract

**Study question:** How does serial sperm donation impact genetic risk in the donor-conceived children and their descendants?

**Summary answer:** In addition to the psychological effects of serial sperm donation, donor-conceived children are at risk of unintentional inbreeding. This risk is compounded by the hard-to-quantify effect of social proximity between mothers. Such inbreeding would cause children to have up to 15% excess risk of childhood mortality or congenital morbidities. The risk to descendants after many generations is spread across many individuals and remains low as long as the number of donor-conceived children does not increase appreciably.

**What is known already:** Inbreeding increases the risk for a range of diseases among the offspring, with the risk increasing with the degree of inbreeding. Sperm donation increases the risk of accidental inbreeding, and thus likely increases disease risk.

**Study design, size, duration:** We performed a literature review of risks associated with consanguinity across a range of traits, together with a model-based mathematical analysis to estimate the short- and long-term risk associated with serial sperm donation.

**Participants/materials, setting, methods:** We used whole-genome sequencing and imputed sequence data from the CARTaGENE longitudinal study to estimate population prevalence of relevant risk alleles. We performed mathematical modelling based on these results on published estimates of the risk associated with inbreeding.

**Main results and the role of chance:** With over 600 children conceived in this serial sperm donation event, 0.1 consanguineous unions would be expected under the simplest model of random mating by generation within the province of Quebec. Preferential mating due to geographic and social proximity among the mothers could increase this rate appreciably, so that accidental inbreeding is not unlikely. Since the likelihood of inbreeding events increases quadratically with the number of children, active inbreeding avoidance by the offspring and interventions to reduce continued serial donation can reduce risk. Over generations, more distant inbreeding is unavoidable, but inbreeding coefficients are reduced. Our model predicts that the long-term excess number of serious adverse events will be fewer than one per generation.

The short- and long-term rates of specific diseases may be affected, however, given public information about the donor carrier status, we expect an excess of 0.84 children per generation [95% CI: 0,3] affected by Hereditary Tyrosinemia of type 1.

**Large scale data:** CARTaGENE is a biobank based in Quebec, Canada, that is accessible following an independent data access protocol and can be found at: https://cartagene.qc.ca/en/

**Limitations, reasons for caution:** Our analysis relies on uncertain estimates of the burden associated with inbreeding. We also rely on simplifying assumptions about future events, including migrations, social interactions between mothers, and future sperm donation events. As a result, our estimates should be seen as coarse estimates.

**Wider implications of the findings:** Serial sperm donation is not uncommon. Each documented instance has raised questions about the genetic burden associated with the practice. By quantifying this risk, this study will help inform the public health and genetic counselling response to these situations, in addition to being of interest from a population genetics perspective.

**Study funding/competing interest(s):** This research was supported by the Canadian Institute for Health Research (CIHR) project grant 437576, NSERC grant RGPIN-2017-04816, the Canada Research Chair program to S.G., and the Canada Foundation for Innovation. T.M.Z was supported by the QLS Grad and Grad Excellence Award. The authors report no competing interests.

**Consanguinity:** The degree of relatedness between individuals, as measured by inheritance from recent ancestors. For example, second cousins share on average 3.125% of their DNA from their great-grandparents.

**Inbreeding:** The production of offspring from individuals with high consanguinity.

**Runs of Homozygosity (ROH):** Stretches of the genome where identical alleles were received from both parents. The fraction of the genome in ROH is a measure of inbreeding.

**Donor-Conceived Child (DCC):** Child born following sperm donation. DCC(X) refers to a child born following sperm donation by individual X.

**Congenital Morbidity:** Diseases or medical conditions present from birth, including physical, intellectual, or developmental. Specifically, does not include any diseases or conditions that arise from exposure to medications or chemicals during gestation or infections during pregnancy.

## Introduction

Recently, documentaries presented instances of serial sperm donation, that is, situations where hundreds of individuals share a single sperm donor.^1,2^ The practice raises many psychosocial, ethical, legal, and genetic questions. For example, what is the genetic risk of unintentional inbreeding in the next generation and the more distant future? How much will this affect the overall population risk? How does the carrier status of the donor inform our interpretation of risk?

While many studies have investigated the genetics and sociology of sperm donation — both regulated and unregulated^3–9^ — we are not aware of formal analyses of the consequences of serial sperm donation.

In this work, we seek to evaluate the genetic implications of serial sperm donation. Since accidental consanguinity is a central concern, we review the literature about the health consequences of consanguineous unions on their offspring, discuss the likelihood of accidental consanguineous unions, the increased risk of specific autosomal recessive diseases, and the long-term genetic consequences.

To illustrate the risk, we will use a recent instance of serial sperm donation in Quebec, Canada, in which three related men (labelled X, Y, and Z in the documentary “*P*è*re 100 Enfants*”) have donated sperm leading to the birth of over 600 donor-conceived children (DCC) since 2008 across the province of Quebec, Canada (see Figure 1).^1^

**Figure 1:**
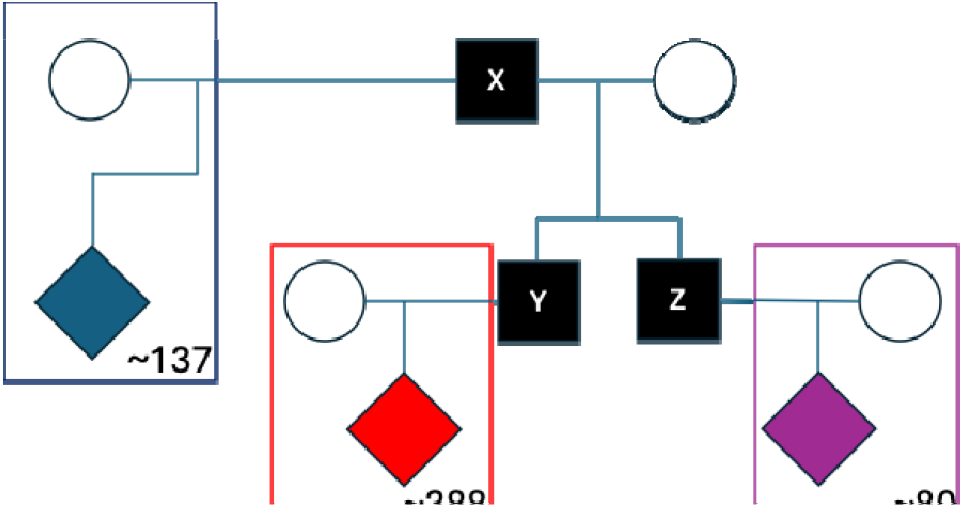
An approximate pedigree describing the serial sperm donation.Following plate notation, the number in the bottom right of the square indicates the number of children associated with each donor reported in a documentary^1^, which we will use in calculations throughout. Many DCCs may not have been included in this report, such that these numbers are likely underestimates. This figure also does not reflect the fact that some mothers had multiple offspring from the donors.

### Consequences of close consanguinity and inbreeding

Consanguinity is associated with offspring mortality and morbidity^10–14^. However, estimates of this contribution vary, and most studies focus on specific outcomes at specific ages. We first review the literature to obtain a coarse estimate of the cumulative consequences, over the life of children, of consanguineous unions.

Global estimates suggest that 10% of the population has parents who are second cousins or closer.^10–12^ To explain this prevalence, consanguineous unions have been hypothesized to be associated with socio-economic benefits such as social cohesion, preservation of family wealth, and maintenance of geographic proximity, that offset some of the genetic drawbacks.^10^ Of course, unintentional consanguinity would not convey any such benefits. By contrast, we expect that there will be many other psychosocial consequences of unintentional consanguinity, including worries about health or possible stigmatization by others. These may be important, but we will not seek to quantify them.

Geneticists have made many attempts at quantifying the impact of consanguineous unions on multiple measures of health, including mortality, congenital morbidity, general complex disease morbidity and, more specifically, infertility.^10,11,15,16^

One of the first cohort studies on under-18 child mortality estimated an excess mortality of 4.8 percentage points in children of first cousins in comparison to children of unrelated individuals.^15,16^ More recently, a meta-analysis determined that, on average, children of first-cousins (with 12.5% autozygosity) will have an excess risk of mortality of 3.5 percentage points after accounting for common confounders such as the negative correlation between consanguinity levels and socioeconomic status.^10^ This meta analysis combined statistics from populations around the world, from 1958 to 2004, and these studies differed in the specific mortality tested, ranging from the rate of stillbirth, the rate of mortality under the ages of 1, 2, and up to 18. There is high variability in reports, with estimates ranging from close to zero excess risk in the stillbirth rate in Turkey to as stark as a 27 excess percentage points of under-18 mortality in South India.^17,18^ A 2020 paper showed that across Bangladesh, there is an excess risk of 10 percentage points in the mortality of children under the age of 5 for children of first cousins.^13^ Social correlates of inbreeding practices complicate the interpretation of these results, and studies vary in methodology to account for these. Mortality rates are also expected to vary based on available medical care^10,11,16^. Given this uncertainty, a conservatively low estimate for the case study, given the availability of healthcare in Quebec and the low overall child mortality, would be 3.5 extra percentage points for children of first-cousin unions.

In addition to mortality, a few morbidities have been associated with the degree of consanguinity. Morbidity itself can be separated into morbidity due to congenital diseases, which primarily affects children, and morbidity due to more complex diseases such as heart disease and breast cancer that might have later onset and lower impact on fitness. Congenital disease morbidity has been shown to increase with consanguinity, with rates dependent on disease: some conditions, such as polydactyly and hydrocephalus, have an overall increased risk of occurring in first-cousin offspring, while other morbidities show population-specific increases due to varying carrier rates for recessive variants, while many morbidities showed no clear association.^19^ Anwar et al. combined morbidities thought to be genetic diseases, and found a 2% overall morbidity among offspring of non-consanguineous families and a 6% overall morbidity among offspring from consanguineous families (second cousins or closer), an excess risk of 4 percentage points.^13^ This estimate only consists of diseases that were either diagnosed in affected individuals or reported in pedigree analysis for affected individuals, but the authors mention that a molecular diagnostic analysis was not conducted.^13^ Some diseases may therefore have been underreported or underdiagnosed. The study that reported no significant difference in stillbirth rates between consanguineous and non-consanguineous in Turkey did find an excess risk of 7.3 percentage points in congenital malformations.^18^ We will use the four excess percentage points from Anwar et al. ^13^ as a conservatively low estimate for the excess congenital morbidity for the offspring of first-cousin unions.

The inbreeding effect on complex diseases and traits has received less attention, and existing studies are sometimes contradictory^10,12,14,16,19–21^. Many studies on multiple complex traits use runs of homozygosity as a measure of consanguinity. Height, weight, and forced expiratory volume were found to be reduced with increased homozygosity^14^ : individuals with a similar degree of homozygosity as the offspring of first-degree relatives were predicted to be on average 3.4 cm shorter than individuals with no runs of homozygosity.^14^ While this research suggests broad impacts of inbreeding on health measures, there is a concern that such multitrait analyses are at least in part confounded by socio-economic factors. Given this, the continuous nature of many complex traits, and the lack of a clear preferred value for many traits (e.g., height), we did not quantify a global excess risk for complex traits that is attributable to consanguineous unions.

One complex trait that has been studied in more detail is fertility. Fertility of consanguineous couples has not been found to be significantly increased or lowered, however, numerous socio-economic confounders exist.^12,22–25^ The fertility of the offspring of a consanguineous marriage has also been studied, with conflicting evidence. An IVF clinic in Lebanon reported that men from consanguineous unions were more likely to be patients who had azoospermia or severe oligospermia.^23^ However, a study in rural Switzerland only reported a negative correlation with maternal inbreeding and family size. They did not find any significant associations between paternal inbreeding and family size.^24^ This finding conflicts with a report by Robert et al., who investigated a population from the Saguenay–Lac-Saint-Jean (SLSJ) region of Quebec, Canada.^26^ The authors found instead that paternal inbreeding had a strong effect on the number of children the couple had, the probability of having children, and the interval between children.^26^ This finding was corroborated by Gunfridsson and Vikström, who examined a historic Swedish population and found that there was only a significant decrease in fertility in inbred men and not inbred women.^25^ The authors found that the more inbred the father, the larger the time interval between children. Finally, a study investigating the UK Biobank found that individuals who carried as much homozygosity as the offspring of first cousins reported a 2.6 times higher chance of being infertile, although this trait did not reach Bonferroni-corrected significance.^14^ Thus, the effect of inbreeding on fertility remains debated.

Both close and distant inbreeding studies suggest a linear relationship between the degree of inbreeding and a range of traits.^10,13,14,16,27,28^ Hence, the doubling in shared DNA (say, between first cousins and half-siblings) would result in a similar doubling of excess risk of mortality, morbidity, and infertility.^13,14,27,28^ Under this assumption, we would estimate that the children from half-sibling unions would have an approximate 7 excess percentage points for childhood mortality, 8 excess percentage points for congenital morbidities, and a difficult-to-quantify but likely significant impact on complex traits.

### Likelihood of unions between related DCC

The public health risk posed by serial sperm donation depends not only on the consequences of inbreeding, but also on the number of unions between DCC related through donors. To coarsely estimate the chance that any one of the serial sperm donors’ children will unintentionally have a child with one of their biological relatives, we must make a set of assumptions. We assume that all the DCC will remain in Quebec, that the mothers of all DCC are distinct and not particularly genetically related to each other, and that the children are monogamous. We will also describe two scenarios, a simplistic baseline where the children conceived from the three sperm donors randomly choose a partner in the overall population of Quebec, and another where we assume increased odds that they meet and have children with each other.

In the first scenario, we can calculate the chance that any half-sibling will have a child with another half-sibling, assuming that they are randomly distributed across Quebec. Suppose that a serial sperm donor *W* has contributed to conceiving *kw* children, evenly split between males and females, and that these children pick partners at random in a population of size *N*. To account for geographic or social proximity between DCCs, we suppose that they have a mating probability increased by a factor s relative to random mating. Then each of the *k_w_*/2 female DCC(W) has a 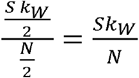 chance of picking a DCC(W) as a partner, so that the overall expected number of such unions is 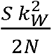 As of 2024, the DCCs are in an age range of 0-16 years old, and we will suppose that they pick partners within the same age group.^29^ There are an estimated 1.4 million children aged 0-15 in the province of Quebec, according to the 2021 Census, so we will assign a population size of *N* = 1.4 million^30^. Since there are 3 sperm donor individuals (Figure 1), the expected number of unions between the respective half-brothers and half-sisters is: 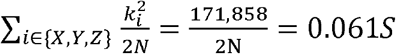. So approximately 0.061s unions between half-siblings at the population level per generation.

In addition, because of the relatedness between the three donors, we expect to find half-avuncular relationships (half-uncle – niece, and half-aunt – nephew) between the DCC with donors × and Y. The expected number of inbreeding events would be: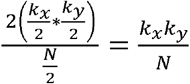. Accounting as well for DCC with donors × and Z, the expected number of avuncular inbreeding events will be: 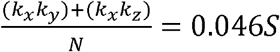 in the first generation. Finally, in this scenario, the number of first-cousin unions between the DCC-conceived children with donors Y and Z is expected as: 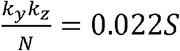.

In total, the expected number of consanguineous unions between the children of the first generation is estimated to be around 0.129s.

Of course, human mate selection is not random. Many factors, including geography, cultural and socio-demographic factors, have been associated with non-random mating. In particular, many of the families are single-parent or LGBTQ+ households and share a history of assisted reproduction. There are many reasons why their children may meet more frequently than expected by chance^31^. We did not find data on assortative mating with respect to parental LGBTQ+ status. If we were to arbitrarily assume that social proximity leads to *s*=10, there would be, on average, 1.3 consanguineous unions between the first generation of children. Of course, the expected number of social interactions or sexual encounters between DCCs would be appreciably higher than the number of consanguineous unions leading to births.

The number of expected consanguineous unions scales quadratically with the number of children. If there were double the number of children than our current estimates (increasing *k* to 1,200 children), the expected number of consanguineous unions would increase fourfold. Conversely, if the children actively avoid consanguineous unions by checking for shared paternity, then the number of expected pairings would decrease. If half of the children actively and successfully avoided consanguineous unions (reducing the at-risk population to 300 children), the expected rate of consanguineous unions would correspondingly decrease fourfold.

### Increased risk of Autosomal Recessive diseases

Based on publicly available information, at least donor × is a carrier of an autosomal recessive (AR) disease, Hereditary Tyrosinemia Type 1 (HT1).^32^ HT1 is a disease classified as an inborn error of metabolism, where the body cannot produce the enzyme FAH, an enzyme found in the metabolic pathway of tyrosine and phenylalanine degradation.^32–34^

In 1992, the drug nitisinone was found to block the faulty pathway upstream of FAH, thereby preventing the build-up of toxic metabolites.^34^ The use of nitisinone in parallel with a low-protein diet has allowed patients with HT1 to live a full and healthy life.^32–34^ Given its relatively high prevalence in Quebec, HT1 has been included in the Quebec newborn screening program since 1970, with a high participation rate.^32^

Any offspring of a heterozygote carrier has a 50% chance of inheriting a copy of the deficient variant from the carrier. This means donors Y and Z are also at risk of being carriers. The offspring will only develop HT1 if they also inherit a copy of a deficient variant (the same or a different deficient variant) from the other parent, since it is an autosomal recessive disease.^33^ In the scenario described in Quebec, the children conceived from sperm donor carriers of HT1 are at risk of developing HT1 only if their mother is also a carrier of HT1.

Through a series of founder events dating back to the colonization of the province of Quebec by France in the 1700s, the regions of Charlevoix, Haute-Côte-Nord and SLSJ and Quebec as a whole have an overrepresentation of HT1 carriers.^35,36^ Previous studies identified more than 98 putative causal mutations in the *FAH* gene and estimated the frequency of overall carriers in the SLSJ region at 1/20.^32,36^ Through recently released whole genome sequencing and imputation conducted by the CARTaGENE biobank (CaG), we found four variants in the gene *FAH* responsible for HT1.^37^ These four variants are classified as “Pathogenic” or “Likely pathogenic” by ClinVar with two stars and no conflicts or interpretation (last accessed on January 26, 2025).^38,39^ To obtain a new estimate of carrier frequencies in Quebec, we imputed the variants on a panel of over 27,000 individuals in CaG using a method described in Mejia-Garcia et al.^37^

We estimate that the cumulative carrier frequency of HT1 carriers in the SLSJ region closer to 1/15 than 1/20, perhaps due to fine-scale geographic differences in sampling between CARTaGENE and the samples used by De Braekeleer et al^36^. The cumulative carrier frequency of known pathogenic or likely pathogenic diseaseausing variants in *FAH* is around 1/110 across Quebec in this cohort (Table 1).

**Table 1:** The four variants in the gene FAH, which have been previously linked to HT1. An estimated carrier frequency was calculated by comparing the number of carriers to the overall population of the CaG. For more information, refer to Table S1.

| Chr | Pos<br>(GRCh38) | Protein/Sequence<br>Variant | ClinVar Classification | Estimated Carrier Frequency |  |
| --- | --- | --- | --- | --- | --- |
|  |  |  |  | Quebec<br>(n=26,738) | SLSJ<br>(n=2,599) |
| 15 | 80181069 | p.Glu364Ter | Pathogenic | 1/1544 | 0 |
| 15 | 80153101 | p.Asn16Ile | Pathogenic or Likely<br>Pathogenic | 1/1275 | 0 |
| 15 | 80168317 | c.606+1G>A | Likely Pathogenic | 1/4889 | 1/519 |
| 15 | 80180230 | c.1062+5G>A | Pathogenic | 1/134 | 1/15 |

As previously mentioned, donor × has been reported to be a carrier of an HT1 risk variant^29^. Treating the status of Y and Z as unknown (with 1/2 carrier probability), the expected number of carriers among the children is 186 (See derivation in Appendix A). Assuming an overall carrier frequency of 1 in 110 in the mothers and uniform distribution across Quebec, we would expect that an average of 1.68 children have HT1 (95% CI [0,3], see Appendix A). The expected number of affected children could be higher or lower depending on the carrier status of Y and Z, and the geographic distributions of the mothers.

The presence of one recessive disease among the children does not indicate that the children are more at risk for genetic diseases overall, since most humans are unknowing carriers for recessive diseases. The uniquely large biological sibship made it easier to identify the carrier status of HT1 and that of other genetic conditions. The increase in the prevalence of HT1 among the children may be compensated for by a reduced risk of being a carrier for other diseases, which have not been identified in the sibship.

### Long-term consequences of serial sperm donation

If we were to assume that each of the children has on average 2 children, then after 2 generations (in approximately 40 to 50 years), there will be approximately 2,400 descendants of these three serial sperm donors. At this point, the most related any descendant in the third generation would be to another descendant in the same generation would be as 2^nd^ cousins sharing an average of 3.125% of their DNA, and most would be half 2^nd^ cousins or more distantly related, sharing less than 2% of their DNA with each other. A common threshold for consanguinity is the degree of 2^nd^ cousins. There is no consensus on the effect of more distant inbreeding on health and fitness^10,11,13,22^. As mentioned above, a parsimonious model is that deleterious effects due to inbreeding are approximately linear in the amount of homozygosity (i.e., identical DNA received from a recent ancestor).

To estimate the future effect of inbreeding caused by serial sperm donation under this model, we therefore compute the probability *P*(*I*) that a future individual *W* is autozygous at a given genetic position because it inherited two copies of the same allele from X. By conditioning on the possible paths linking w to X, and under random mating assumptions across Quebec, we find *P*(*I*) ≃ 1.72 × 10-8 in one non-overlapping generation (Appendix B). While this random mating is obviously a simplification, we feel that it provides a conservatively high estimate, in that any interprovincial or international migration would reduce *P*(*I*), and levels of interregional migrations would have to be reduced drastically from the current levels to maintain fine-scale population structure over tens of generations. Given that the human genome is approximately 3 billion base pairs – this means that on average, 52 bases will be autozygous between two distant relatives of X, Y, and Z. Considering that the offspring of first cousins have a 1/16 chance of autozygosity and a 7% increase of congenital morbidity, we expect a very small risk per individual (close to 1.7 in 100 million). In a population size of 9 million, giving an effective per-generation N of 1.4 million, the number of child mortality or congenital morbidity events would be less than one individual per 100 generations.

## Conclusion

The presence of serial sperm donors appears to be a recurrent feature of unregulated sperm donation. In this study, we focused on the short- and long-term genetic consequences of serial sperm donation and its impact on the frequency of tyrosinemia variants across Quebec. While we focused on genetic consequences, serial sperm donations have important legal, ethical, social and psychological consequences. We did not cover these here, primarily because they are outside our area of expertise. While the short-term consequences of unintentional consanguinity can be severe, the likelihood of unintentional consanguinity can be reduced through awareness and communication of paternity. On the contrary, the long-term consequences of serial sperm donation depend on the choices of many individuals. The long-term risk at the population scale remains low unless many additional donations are made by the serial donors or their relatives.

Given a population of 8.8 million and a carrier frequency of 1/110, there are approximately 80,000 HT1 carriers in Quebec. The extra 260 carriers do not qualitatively change the frequency of HT1 carriers in Quebec, yet they would lead to two additional cases per generation even under random mating. There is no evidence that this increase would be accompanied by an overall increase in the recessive genetic load – the slight increase in HT1 burden may be compensated by a minute decrease in the frequency of many recessive alleles not carried by X, Y, and Z. Given the efficient newborn screening program already in place, we expect that most if not all affected children will receive the available treatment ^32^.

In conclusion, health professionals involved in the care of the affected mothers and children can discuss measures to prevent short-term accidental inbreeding, such as awareness and communication of paternity, while reassuring them if they have any concerns about the long-term consequences for their descendants. Counselling for the children regarding HT1 status should be in line with anyone with a biological parent who is a carrier.

## Methods

Estimates of the carrier frequencies were conducted following the approach described in Mejia-Garcia et al^37^.

Confidence intervals for the estimation of carriers were computed as a convolution of four equally weighted Poisson distributions as described in Appendix A.

## Data availability

The data used for this project is from the CARTaGENE biobank. The data is accessible upon request and can be found at:https://cartagene.qc.ca/.

## Contributions

Conceptualization: T.M.Z. and S.G., Methodology: T.M.Z., A.M-G, S.G., Analysis: T.M.Z., A.M-G, S.G., Writing – Original Draft: T.M.Z., A. M-G, S.G., Interpretation of the results: T.M.Z., C.B., C. L., A.M.L, S.G., Writing – Review and Editing: T.M.Z., C.B., C. L., A.M.L, S.G.

## Acknowledgements

We would like to thank the mothers that we met with for sharing their useful perspectives. We would also like to thank Bailey Irene Midori Hoy for her translation work.

## Appendix A Expected number of Tyrosinemia cases among children

Assuming that × is an HT1 heterozygote carrier, there are four possible sets D of carriers among the donors, *D* ∈ {{*X*}, {*X, Y* }, {*X, Z* }, {*X, Y, Z* }}.

Each scenario has a probability of 0.25 due to Mendelian segregation. If donor *i* has *ki* children, the expectation for the number c of children who inherited a risk allele from a donor is: 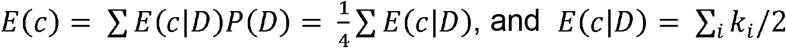.

We can thus compute

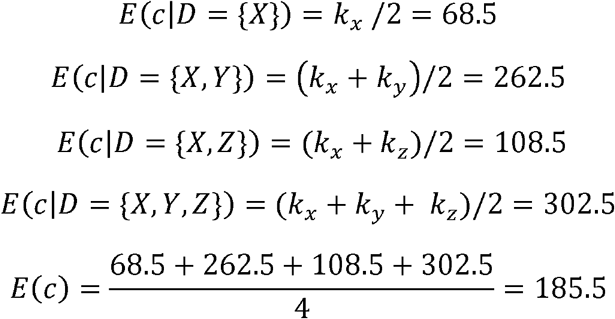

Therefore, the expected number of carriers of HT1 solely due to their donor is 186, 95% CI: [64,313], with most of the variance coming from *D*. Assuming an overall carrier frequency of 1 in 110 in the mothers and uniform distribution across Quebec, the expectation is that 0.84 children will have HT1, with 95% CI: [0,3].

## Appendix B Relatedness among DCCs and long-term autozygosity.

DCCs in our case study have many half-siblings, half-uncles/aunts, and first cousins. For pairs of children with distinct and unrelated mothers, a DCC with donor × (or DCC(X)) would, on average, share 25% of their DNA with another DCC(X) and 12.5% of their DNA with DCC(Y) and DCC(Z). Similarly, a DCC(Y) would share 25% of their DNA with another DCC(Y) and 12.5% of their DNA with a DCC(X) – their half-uncle/aunt – and 12.5% of their DNA with a DCC(Z), their first cousin (See Figure 1).

Let *W* be an individual living in the distant future. We would like to compute the probability *P*(*I*) that this individual will be autozygous at a given genetic position due to serial sperm donation. We can condition on the probability that the alleles received by *W* came from the DCCs as: *P*(*I*) = ∑ *P*(*I*|*ij*)*P*(*ij*), where *ij* represents the event that *W*inherited alleles from the DCC(*i*) and DCC(*j*), with *i, j* ∈ {*X, Y, Z* }

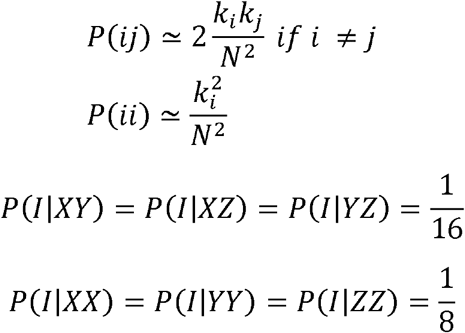

Combining it all, we have 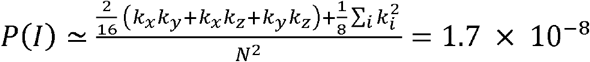.

To estimate genetic consequences on health, we assume a linear relationship between autozygosity and the percent increase chance of morbidity. Since the offspring of first cousins have a 6.25% probability of autozygosity and an increased morbidity of 7.5 percentage points of mortality and congenital disease, we have approximately 1 percentage point of load per percentage point in autozygosity. We can assign an approximate increased risk of 1.7/100,000,000 per generation for descendants of X.

---

**Figure A1:**
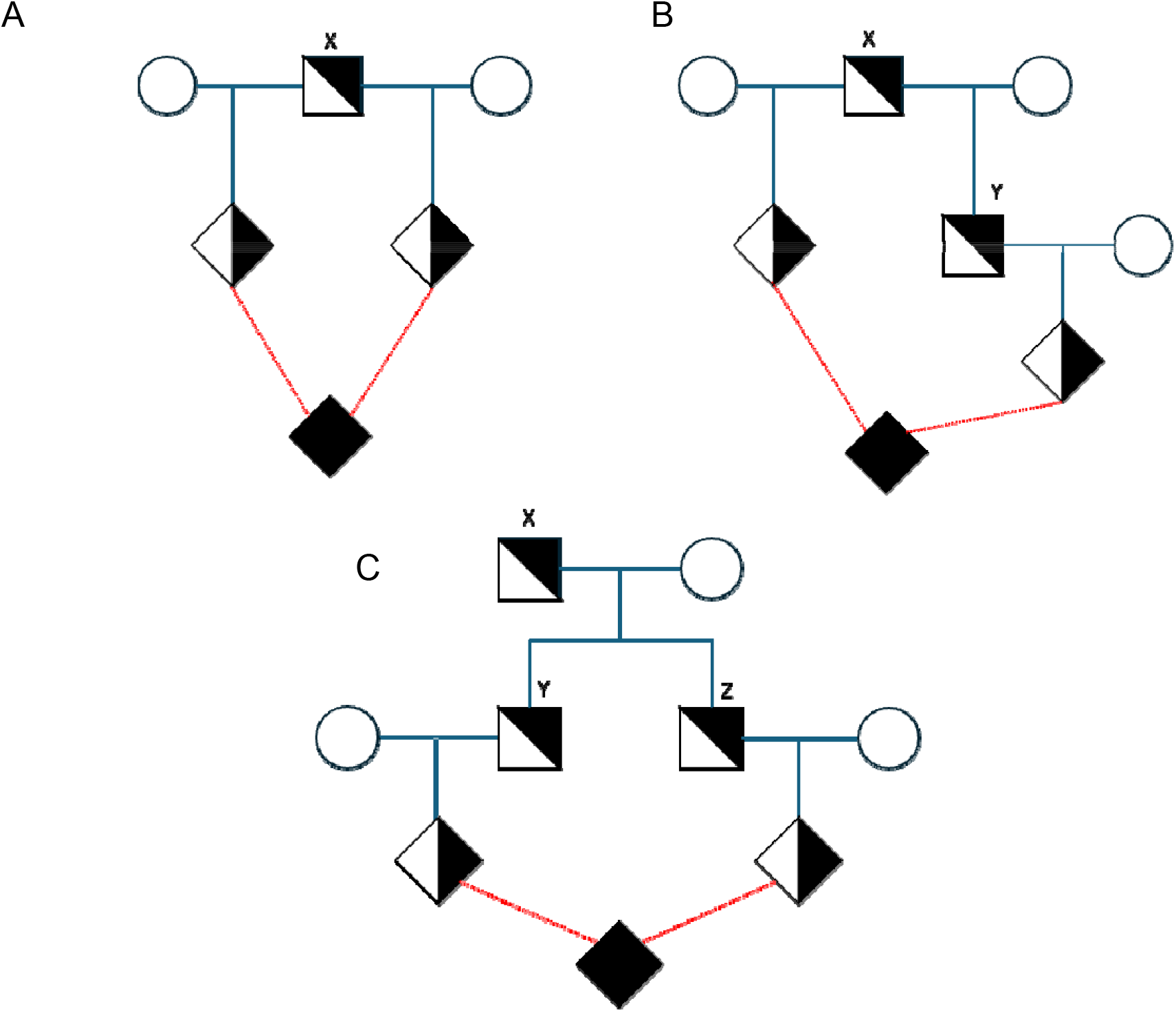
The probability that any future offspring of X, Y, and Z is autozygous given that they inherited genetic information from the donors. To calculate the chance that any single position of the genome will be autozygous, we determine the likelihood of transmission to the descendant. In **Fig. A1A**, we show the probability that a descendant from the same parent (XX, YY, or ZZ) will be autozygous. For any position in the genome, there is a ½ chance that the donor will pass on a given SNV to one child, and another ½ chance that the donor will pass the same SNV onto another child. If these two children, then have a descendant together, given that are passing on their genetic material, then they will each have a ½ chance of passing the same SNV to the F_2_ child leading to a (1/2)^4^ chance of autozygosity for that given position, however since × is diploid, there is an equal, disjoint chance that the F_2_ child is autozygous for the other chromosome, leading to a final chance of **Fig. A1B** shows the probability of autozygosity when × and Y are the ancestors, this will also be the same for × and Z. Because × and Y only share 50% of their DNA, there is a (½)^2^ chance that they will pass on the same SNV to their children. From there, each child individually has a ½ chance to pass on the same SNV to the F_2_ child, which means that any descendant of × and Y will have a (½)^4^ chance to be autozygous in any given position. **Fig. A1C** shows the probability of autozygosity when Y and Z are the ancestors. Note that the probability is the same as × and Y, and × and Z, because we are considering that Y and Z are full siblings. Y and Z share 50% of their DNA, and as such, there is a (½)^2^ chance that they will pass on the same SNV to their children. Each child has a 50% chance of passing on the same SNV to the affected child, resulting in a (½)^4^ chance for the affected child to be autozygous in a given position.

**Table S1:** The four variants in the gene FAH, which have been previously linked to HT1.We report the Chromosome, Position in hg38, Reference, Alternate, and the Protein or Sequence ID. Additionally, we report ClinVar’s Classification, the number of carriers in the CaG separated by both whole genome sequencing and after imputation. Finally, using the estimated samples from both Quebec and SLSJ, we calculated the estimated allele frequency. The ClinVar classification P stands for Pathogenic, LP = Likely Pathogenic.

| Chr | Position<br>(GRCh38) | Reference | Alternate | Protein/<br>Sequence ID | ClinVar<br>Classification | Number of Carriers<br>in CaG |  | Estimated Allele<br>Frequency |  |
| --- | --- | --- | --- | --- | --- | --- | --- | --- | --- |
|  |  |  |  |  |  | WGS<br>(n =<br>2173) | Imputed<br>(n =<br>29,337) | Quebec<br>(n =<br>26,738) | SLSJ (n =<br>2599) |
| 15 | 80181069 | G | T | p.Glu364Ter | P | 2 | 19 | 1/1544 | 0 |
| 15 | 80153101 | A | T | p.Asn16Ile | P/LP | 1 | 23 | 1/1275 | 0 |
| 15 | 80168317 | G | A | c.606+1G>A | LP | 1 | 6 | 1/4889 | 1/519 |
| 15 | 80180230 | G | A | c.1062+5G>A | P | 19 | 219 | 1/134 | 1/16 |

## References

1. Pere 100 enfants. All Episodes (2023).

2. The Man with 1000 Kids. (2024).

3. Donovan, C. Genetics, Fathers and Families: Exploring the Implications of Changing the Law in Favour of Identifying Sperm Donors. Soc. Leg. Stud. 15, 494–510 (2006).

4. Gong, D., Liu, Y.-L., Zheng, Z., Tian, Y.-F. & Li, Z. An overview on ethical issues about sperm donation. Asian J. Androl. 11, 645–652 (2009).

5. Bauer, T. & Meier-Credner, A. Circumstances Leading To Finding Out about Being Donor-Conceived and Its Perceived Impact on Family Relationships: A Survey of Adults Conceived via Anonymous Donor Insemination in Germany. Soc. Sci. 12, 155 (2023).

6. Serre, J.-L. et al. Does anonymous sperm donation increase the risk for unions between relatives and the incidence of autosomal recessive diseases due to consanguinity? Hum. Reprod. 29, 394–399 (2014).

7. Maron, J. L. A Hidden Generation: Offspring from Sperm Donation in an Era of Medical Paternalism. Clin. Ther. 42, 2119–2121 (2020).

8. Pennings, G. A SWOT analysis of unregulated sperm donation. Reprod. Biomed. Online 46, 203–209 (2023).

9. Marvel, S. “Tony Danza Is My Sperm Donor?”: Queer Kinship and the Impact of Canadian Regulations around Sperm Donation. Can. J. Women Law 25, 221–248 (2013).

10. Bittles, A. H. & Black, M. L. Consanguinity, human evolution, and complex diseases. Proc. Natl. Acad. Sci. 107, 1779–1786 (2010).

11. Bittles, A. H. A Community Genetics Perspective on Consanguineous Marriage. Public Health Genomics 11, 324–330 (2008).

12. Jaber, L., Nashif, A. & Diamond, G. Consanguinity, Fertility and Reproductive Outcomes: An International Review. Med. Res. Arch. 11, (2023).

13. Anwar, S., Taslem Mourosi, J., Arafat, Y. & Hosen, M. J. Genetic and reproductive consequences of consanguineous marriage in Bangladesh. PLOS ONE 15, e0241610 (2020).

14. Clark, D. W. et al. Associations of autozygosity with a broad range of human phenotypes. Nat. Commun. 10, 4957 (2019).

15. Neel, J. V. & Schull, W. J. The Effect of Inbreeding on Mortality and Morbidity in Two Japanese Cities. Proc. Natl. Acad. Sci. U. S. A. 48, 573–582 (1962).

16. Schull, W. J. & Neel, J. V. The effects of parental consanguinity and inbreeding in Hirado, Japan. V. Summary and interpretation. Am. J. Hum. Genet. 24, 425–453 (1972).

17. Reddy, P. G. Effects of inbreeding on mortality: a study among three South Indian communities. Hum. Biol. 57, 47–59 (1985).

18. Alper, Ö. M. et al. Consanguineous marriages in the province of Antalya, Turkey. Ann. Génétique 47, 129–138 (2004).

19. Rittler, M., Liascovich, R., López-Camelo, J. & Castilla, E. E. Parental consanguinity in specific types of congenital anomalies. Am. J. Med. Genet. 102, 36–43 (2001).

20. Teeuw, M. E. et al. Do consanguineous parents of a child affected by an autosomal recessive disease have more DNA identical-by-descent than similarly-related parents with healthy offspring? Design of a case-control study. BMC Med. Genet. 11, 113 (2010).

21. Hamamy, H. Consanguineous marriages: Preconception consultation in primary health care settings. J. Community Genet. 3, 185–192 (2012).

22. Helgason, A., Pálsson, S., Guðbjartsson, D. F., Kristjánsson, Þ. & Stefánsson, K. An Association Between the Kinship and Fertility of Human Couples. Science 319, 813–816 (2008).

23. Inhorn, M. C., Kobeissi, L., Nassar, Z., Lakkis, D. & Fakih, M. H. Consanguinity and family clustering of male factor infertility in Lebanon. Fertil. Steril. 91, 1104–1109 (2009).

24. Postma, E., Martini, L. & Martini, P. Inbred women in a small and isolated Swiss village have fewer children. J. Evol. Biol. 23, 1468–1474 (2010).

25. Gunfridsson, E. H. & Vikström, L. Long-term health outcomes from inbreeding in a historical Swedish population: longevity, fertility, and impairments. Ann. Hum. Biol. 51, 2369281 (2024).

26. Robert, A., Toupance, B., Tremblay, M. & Heyer, E. Impact of inbreeding on fertility in a pre-industrial population. Eur. J. Hum. Genet. 17, 673–681 (2009).

27. Malawsky, D. S. et al. Influence of autozygosity on common disease risk across the phenotypic spectrum. Cell 186, 4514-4527.e14 (2023).

28. Fried, K. & Davies, A. M. Some effects on the offspring of uncle-niece marriage in the Moroccan Jewish community in Jerusalem. Am. J. Hum. Genet. 26, 65–72 (1974).

29. Pere 100 enfants. Episode 4 (2023).

30. Statistics Canada. 2021 Census of Population. Statistics Canada Catalogue no. 98-316-X2021001. (2023).

31. Massey, D. J., Szpiech, Z. A. & Goldberg, A. Differentiating mechanism from outcome for ancestry-assortative mating in admixed human populations. Preprint at 10.1101/2024.06.06.597727 (2024).

32. Hereditary Tyrosinemia: Pathogenesis, Screening and Management. vol. 959 (Springer International Publishing, Cham, 2017).

33. Russo, P. A., Mitchell, G. A. & Tanguay, R. M. Tyrosinemia: A Review. Pediatr. Dev. Pathol. 4, 212–221 (2001).

34. Holme, E. & Lindstedt, S. NONTRANSPLANT TREATMENT OF TYROSINEMIA. Clin. Liver Dis. 4, 805–814 (2000).

35. Paradis, K. Tyrosinemia: the Quebec experience. Clin. Investig. Med. Med. Clin. Exp. 19, 311–316 (1996).

36. De Braekeleer, M. & Larochelle, J. Genetic epidemiology of hereditary tyrosinemia in Quebec and in Saguenay-Lac-St-Jean. Am. J. Hum. Genet. 47, 302–307 (1990).

37. Mejia-Garcia, A. et al. Using the ancestral recombination graph to study the history of rare variants in founder populations. Preprint at 10.1101/2025.03.13.643149 (2025).

38. McLaren, W. et al. The Ensembl Variant Effect Predictor. Genome Biol. 17, 122 (2016).

39. Landrum, M. J. et al. ClinVar: public archive of relationships among sequence variation and human phenotype. Nucleic Acids Res. 42, D980–985 (2014).

